# Concordance between amyloid-PET quantification and real-world visual reads: results from IDEAS

**DOI:** 10.1101/2024.10.31.24316518

**Authors:** Ehud Zeltzer, Daniel R. Schonhaut, Nidhi S. Mundada, Ganna Blazhenets, Jhony Mejia Perez, David Soleimani-Meigooni, Hanna Cho, Kamalini Ranasinghe, Charles Windon, Golnaz Yadollahikhales, Charles Apgar, Constantine Gatsonis, Maria C. Carrillo, Lucy Hanna, Justin Romanoff, Bruce E. Hillner, Robert Koeppe, Andrew March, Barry A. Siegel, Karen Smith, Rachel A. Whitmer, Leonardo Iaccarino, Gil D. Rabinovici, Renaud La Joie

## Abstract

Amyloid-PET detects fibrillar β-amyloid deposits, a defining feature of Alzheimer’s disease. This technology has been used in research for over 20 years, and is now used in clinical practice to guide patient diagnosis and management. However, the real-world use of amyloid-PET may differ from research settings due to less standardized acquisition protocols, less experienced visual readers, and patients with more comorbidities, potentially compromising test accuracy. We evaluated the performance of amyloid-PET as used in a real-world clinical setting utilizing data collected via the Imaging Dementia—Evidence for Amyloid Scanning (IDEAS) study.

The study collected clinical amyloid-PET scans of older adults with cognitive decline acquired in 294 imaging facilities using three FDA-approved radiotracers. We centrally processed these scans using a PET-only processing pipeline and derived summary quantification of cerebral radiotracer retention measured on the Centiloid scale. We applied an *a priori* autopsy-based threshold of 24.4 Centiloids to quantitatively define scan positivity and compared this quantitative classification with binary visual reads performed by local radiologists or nuclear medicine physicians.

10,350/10,774 scans (96%) passed quality control and were successfully quantified. Median patient age was 75 (interquartile range 71, 80) years, 51% were females, 87% self-identified as White, 63% had mild cognitive impairment (vs. 37% with dementia), and 61% of scans were visually positive based on local reads. Centiloid values were higher in visually positive scans (median=74 [46, 99]) compared to scans locally read as negative (−2 [−13, 12]; p<0.001). Patients with dementia had higher Centiloids than those with mild cognitive impairment (57 [8, 91] vs. 34 [−2, 79]; p<0.001), consistent with a higher visual positivity rate (70% vs. 56%, respectively; p<0.001). Agreement between local visual reads and quantitative classification was 86.3% (95%CI: 85.7%, 87.0%), corresponding to Cohen’s Kappa of 0.715 (95%CI: 0.701, 0.729; p<0.001). Overall, 5,519 [53% of total] scans were positive by both visual read and quantification (V+/Q+); 3,416 [33%] were negative by both (V-/Q-), 813 [8%] were V+/Q-, and 602 [6%] were V-/Q+. Female sex, White race, and use of the radiotracers ^18^F-flutemetamol and ^18^F-florbetaben (compared to ^18^F-florbetapir) were associated with higher visual-quantitative concordance.

In conclusion, local visual reads showed high concordance with central quantification of clinical amyloid-PET scans, supporting the validity of amyloid-PET as used in the real world and the use of quantification in clinical settings.

## Introduction

Brain amyloid-β (Aβ) deposition is a defining feature of Alzheimer’s disease (AD) neuropathological changes.^1^ Since its development 20 years ago, Aβ-PET has been widely used in AD clinical research and drug development to detect Aβ plaques in vivo.^2^ In the clinical management of patients with cognitive symptoms, Aβ-PET affects aetiologic diagnosis, use of medications, and referral for additional work-up.^3–5^ However, previous research studies that evaluated the validity of Aβ-PET typically included highly selected participants, and used harmonized PET acquisition protocols, quantitative analysis often relying on co-acquired MRI, and central PET interpretation by highly experienced experts. In contrast, the use of Aβ-PET in real-world clinical settings differs from research studies in important respects, including diverse patient populations with more numerous neurological and medical comorbidities, heterogeneous acquisition techniques, and less experienced interpreters, which may compromise the accuracy of the test. Concerns over the generalizability of Aβ-PET research findings to real-world clinical practice have been raised by academia^6–8^ and governmental agencies.^9^

Manufacturer-developed and regulatory agency-approved interpretation methods of Aβ-PET using the clinically approved radiotracers (^18^F-florbetapir,^10^ ^18^F-florbetaben,^11^ and ^18^F-flutemetamol^12^) employ visual inspection by a physician and dichotomous interpretation of the scan as positive or negative for cortical radiotracer retention.^13–15^ Alternatively, quantitative analysis allows the calculation of a standardized uptake value ratio (SUVR) and its conversion to Centiloids (CL), a standardized measure of cerebral radiotracer retention across radiotracers and preprocessing pipelines.^16–20^ The CL scale is, by definition, anchored at an average of 0 for young controls without Aβ, and at 100 for patients with mild-moderate dementia due to AD.^20^ In research, SUVR and CL quantification have been used to define Aβ positivity based on thresholds (typically ranging between 10-40 CL),^21–23^ and high concordance was found between expert visual reads and CL calculated using state-of-the-art PET data and processing pipelines.^24,25^ Similar concordance in the clinical setting would support the validity of the test as used in the real world.

To assess the performance of Aβ-PET in the real world, we analyzed a large sample of clinical scans acquired in patients with mild cognitive impairment (MCI) or dementia enrolled in the Imaging Dementia—Evidence for Amyloid Scanning (IDEAS) Study, a large, real-world study evaluating the clinical utility of Aβ-PET under the Centers for Medicare & Medicaid Service’s Coverage with Evidence Development program. Study sites acquired and visually interpreted PET scans locally and shared the images in a study image archive for central preprocessing and quantification. We aimed to assess the concordance between real-world local visual reads and centrally extracted CL and characterize factors associated with visual-quantitative discordance.

## Methods

### Study oversight and design

The IDEAS study was designed as a single-group, multisite study to assess the clinical utility of Aβ-PET in Medicare beneficiaries with cognitive impairment meeting the 2013 Appropriate Use Criteria (AUC) for Aβ-PET.^26,27^

The study design and procedures were previously described in detail.^4^ In short, the study was managed by the American College of Radiology (ACR) under a central institutional review board (IRB) (Advarra, formerly Schulman Associates), with some sites requiring local IRB approval. Written informed consent for patient participation was obtained by the dementia specialist (see below), either directly from the patient or, in instances in which the specialist determined that the patient lacked the capacity to consent, from a legally authorized representative with patient assent.

### Study population

Patients were recruited by dementia specialists from their clinical practices. Eligible patients were Medicare beneficiaries aged 65 years or older, English- or Spanish-speaking, with a diagnosis of MCI or dementia established by the specialist within the prior 24 months. All patients were required to have completed a comprehensive diagnostic assessment, including global cognition assessed via the Mini-Mental State Examination (MMSE, range 0 [worst] to 30 [best]) or Montreal Cognitive Assessment (MoCA, range 0 [worst] to 30 [best]) at the time of enrollment, laboratory testing within the past 12 months, and head computed tomography or MRI within the past 24 months. Patients were further required to meet AUC for Aβ-PET: (1) the aetiologic cause of cognitive impairment remained uncertain after a comprehensive evaluation by the dementia specialist; (2) AD was a diagnostic consideration; and (3) knowledge of Aβ-PET status was expected to alter diagnosis and management.^26^ Patients were excluded if Aβ status was already known based on prior PET or cerebrospinal fluid analysis, or if learning Aβ status could, in the opinion of the specialist, cause significant psychological harm.

### Clinical evaluation

Before PET acquisition, the dementia specialist completed a pre-PET case report form that described the patient’s demographics and clinical characteristics, including level of impairment (dementia or MCI), MMSE or MoCA scores, and history of vascular risk factors. All clinical data included in the current paper were collected before Aβ-PET acquisition.

### PET acquisition and visual interpretation

Aβ-PET scans were acquired at accredited imaging facilities across the U.S. between February 16, 2016, and January 10, 2018. Scans were completed within 30 days of the pre-PET assessment using any of the three FDA–approved Aβ-PET radiotracers: ^18^F-florbetapir, ^18^F-flutemetamol, and ^18^F-florbetaben. Sites were instructed to acquire scans following FDA labels and published practice guidelines,^28^ but no harmonized protocol was provided.

Scans were interpreted locally at the performing facilities by imaging specialists. The imaging specialists were required to be board certified in diagnostic radiology or nuclear medicine and to have successfully completed vendor-provided, radiotracer-specific training for interpreting Aβ-PET scans. Interpretations followed the approved reading methods of each radiotracer^13–15^ and classified each scan as i) “Positive for cortical Aβ,” ii) “Negative for cortical Aβ”, or iii) “Uninterpretable/technically inadequate study.” Four scans that were classified as uninterpretable/technically inadequate and seven scans missing visual reads were excluded from analyses involving visual reads. Imaging specialists also provided a read confidence level (low, intermediate, or high) and indicated whether a quantification method was used to assist visual interpretation. No information is available on the quantification methods used or procedures for incorporating quantification with the FDA-approved visual reading methods.

### PET quantitative processing

Unless the individual patient or the imaging center opted out, PET facilities submitted images to the ACR IDEAS image archive using the web-based TRIAD^TM^ application, which anonymized the scans. Overall, Aβ-PET data from 10,774 patients were uploaded to the Image and Data Archive (IDA) of the Laboratory of Neuroimaging (LONI) and later downloaded for central processing at the University of California, San Francisco. Image preprocessing, quality control, and quantification were performed between December 2021 and April 2023. Because structural imaging (i.e., head CT or MRI) was unavailable, PET scans were processed using the previously validated robust PET-only processing (rPOP, https://github.com/LeoIacca/rPOP) pipeline, which was specifically designed to analyze community-acquired heterogeneous Aβ-PET data.^29^ CL were extracted from scans that passed quality checks (Supplementary PET processing methods). We excluded 413 scans for failing CL-extraction quality assurance, with the remaining 10,361 scans included in current analyses (Supplementary PET processing methods).

### Quantification-based threshold for Aβ-PET positivity

We applied a CL threshold to dichotomize the quantitative PET data as positive/negative so that quantitative classification could be compared to visual reads. Primary analyses were based on an *a priori*-defined threshold of 24.4 CL derived from an independent, multi-site study evaluating CL values versus AD neuropathological changes (ADNC).^30^ In autopsy-proven cases, a threshold of 24.4 CL optimally distinguished intermediate-high from none-low ADNC with 85.5% overall accuracy (84.1% sensitivity, 87.9% specificity). This threshold also optimally distinguished Thal Aβ Phases 4-5 from Phases 0-3 (87.7% accuracy, 96.4% sensitivity, 78.2% specificity).

Yet, the choice of a specific value to dichotomize CL is relatively arbitrary as various thresholds have been proposed based on different standards of truth and statistical approaches. To reflect this variability and acknowledge the existence of an “intermediate zone” in Aβ-PET values,^31^ we conducted a literature search for CL positivity thresholds (Supplementary Table 1). Based on this search, we operationalized a borderline/intermediate zone as a CL value between 10 and 40 (i.e., about ±15 CL around our *a priori* 24.4 threshold). This category is meant to highlight that scans within this CL range could be classified as positive or negative based on different published thresholds. We then compared visual-quantitative agreement inside and outside this intermediate zone and calculated concordance using thresholds across this range.

### Statistical analysis

Mean differences between groups were compared using Welch’s t-test, and proportional differences using the Chi-Square Test. Because of non-normal distribution, we used Mann-Whitney U test to compare CL across groups. All tests are two-tailed, with p<0.05 considered statistically significant. Effect sizes were reported using Cohen’s D for t-test, rank biserial correlation for Mann-Whitney U test, and Cramer’s V for the Chi-Square Test.

Concordance between visual reads and quantitative classification using the *a priori* 24.4-CL positivity threshold was assessed by calculating: (1) raw percent agreement, and (2) Cohen’s Kappa, which corrects for chance-level agreement. The level of agreement indicated by Cohen’s Kappa was interpreted using thresholds recommended by McHugh.^32^ We tested for statistically significant differences in Cohen’s Kappa as described by Fleiss et. al.^33^ Additionally, we calculated Cohen’s Kappa between visual reads and quantitative classification across a range of CL thresholds, from 0 to 60 CL, in 0.1 CL increments. We then identified the CL thresholds with optimal visual-quantitative concordance according to i) maximal Cohen’s Kappa and ii) maximal receiver operating characteristic curve Youden’s index. A permutation testing procedure was used to assess differences in (1) the strength of visual-quantitative agreement and (2) optimal CL thresholds between quantification- and non-quantification-supported visual reads (Supplementary Methods).

To explore drivers of visual-quantitative discordance, we used logistic regression to predict visual positivity over negativity, first by CL alone, and then adding age, sex, radiotracer, impairment level (MCI/dementia), race, ethnicity, and presence of at least one vascular risk factor or comorbidity (congestive heart failure, atrial fibrillation, ischemic heart disease, hypertension, dyslipidaemia, diabetes mellitus, cerebrovascular disease, or current tobacco use), as covariates. We excluded one case missing covariate data from this analysis. We used the area under the receiver operating characteristic curve (ROC-AUC, based on a non-parametric estimate) to assess these models’ performance and the Chi-squared test to compare them.

To explore the relationship between visual read confidence and CL, we plotted the proportion of scans with low, intermediate, and high confidence ratings as a function of CL, applying an automated Gaussian kernel smoothing using the seaborn package in Python.^34^

Statistical analyses were performed using jamovi (version 2.3, The jamovi project), MATLAB (version R2022b, The MathWorks Inc., Natick, Massachusetts), Python (version 3.10), and SciPy (version 1.11.4)^35^. Graphs were created using GraphPad Prism (version 10, GraphPad Software, San Diego, California) and Microsoft Excel 365 (Microsoft Corporation, Redmond, Washington).

## Results

All 10,361 patients with available CL quantification were included, and their characteristics are presented in Table 1. Patients were evaluated at 500 clinical sites by 782 dementia specialists, and scans were acquired in 294 facilities and visually read locally by 602 imaging specialists. The mean patient age was 76 years, 51% were female, and 68% had above high school education. Most patients self-identified as White (88%), 3% as Black, and 4% as Hispanic. Patients were more commonly diagnosed with MCI (63%) than dementia (37%), and 71% had at least one vascular risk factor or comorbidity. ^18^F-florbetapir was the most commonly used radiotracer (64%), followed by ^18^F-florbetaben (29%) and ^18^F-flutemetamol (6%).

**Table 1.** Patient characteristics.

|  | All<br>(100%) | Visually positive<br>(61%) | Visually negative<br>(39%) | p | ES |
| --- | --- | --- | --- | --- | --- |
| N | 10,361 | 6,332 | 4,018 | <0.001 | NA |
| Age, median (IQR) | 75 (71, 80) | 76 (72, 81) | 74 (70, 79) | <0.001 | 0.122 |
| Female (%) | 5,245 (51) | 3,317 (52) | 1,924 (48) | <0.001 | 0.044 |
| **Education** |  |  |  |  |  |
| No formal education (%) | 57 (0.6) | 25 (0.4) | 32 (0.8) |  |  |
| Grade school (%) | 228 (2.2) | 127 (2.0) | 101 (2.5) |  |  |
| Some high school (%) | 411 (4.0) | 249 (3.9) | 162 (4.0) |  |  |
| High school (including equivalency, %) | 2,621 (25.3) | 1,642 (25.9) | 978 (24.3) | <0.001 | 0.056 |
| Some college or associate degree (%) | 2,492 (24.1) | 1,464 (23.1) | 1,025 (25.5) |  |  |
| Bachelor's degree (%) | 2,415 (23.3) | 1,520 (24.0) | 892 (22.2) |  |  |
| Master's degree (%) | 1,274 (12.3) | 813 (12.8%) | 458 (11.4) |  |  |
| Doctorate (%) | 863 (8.3) | 492 (7.8%) | 370 (9.2) |  |  |
| **Race and ethnicity** |  |  |  |  |  |
| American Indian (%) | 23 (0.2) | 11 (0.2) | 12 (0.3) | 0.19 | 0.013 |
| Alaskan Native (%) | 1 (0.0) | 1 (0.0) | 0 (0.0) | 0.43 | 0.008 |
| Asian (%) | 188 (1.8) | 88 (1.4) | 100 (2.5) | <0.001 | 0.040 |
| Black (%) | 316 (3.1) | 174 (2.7) | 142 (3.5) | 0.023 | 0.022 |
| Native Hawaiian or Pacific Islander (%) | 8 (0.1) | 3 (0.0) | 5 (0.1) | 0.17 | 0.014 |
| White (%) | 9,125 (88.2) | 5,690 (89.9) | 3,435 (85.5) | <0.001 | 0.066 |
| **Ethnicity** |  |  |  |  |  |
| Hispanic or Latino (%) | 449 (4.3) | 251 (4.0) | 198 (4.9) | 0.019 | 0.023 |
| **Past medical history** |  |  |  |  |  |
| Vascular Risk Factor/Comorbidity<sup>a</sup> (%) | 7,323 (71) | 4,453 (70) | 2,860 (71) | 0.35 | 0.009 |
| **Impairment level** |  |  |  |  |  |
| Dementia (%) | 3,861 (37) | 2,701 (43) | 1,154 (29) | <0.001 | 0.140 |
| MCI (%) | 6,500 (63) | 3,631 (57) | 2,864 (71) |  |  |
| MMSE, median (IQR) | 26 (22, 28)<br>(n=8,114) | 25 (21, 27)<br>(n=4,883) | 27 (24, 28)<br>(n=3,223) | <0.001 | 0.243 |
| MoCA, median (IQR) | 22 (18, 25)<br>(n=2,728) | 21 (17, 24)<br>(n=1,753) | 23 (20, 26)<br>(n=973) | <0.001 | 0.273 |
| **Tracer** |  |  |  |  |  |
| <sup>18</sup>F-florbetapir | 6,699 (65) | 4,078 (64) | 2,612 (65) |  |  |
| <sup>18</sup>F-florbetaben | 3,033 (29) | 1,843 (29) | 1,189 (30) | 0.076 | 0.022 |
| <sup>18</sup>F-flutemetamol | 629 (6) | 411 (7) | 217 (5) |  |  |

Demographic and clinical characteristics of patients whose Aβ-PET scans were retained for analysis. P-values and effect sizes reflect differences between visually positive and visually negative scans. ES denotes rank biserial correlation and Cramer's V for continuous and categorical variables, respectively. Four scans that were classified as uninterpretable or technically inadequate and seven scans missing visual reads are included in the "All" column only. ES = effect size; IQR = Interquartile range; MCI = Mild cognitive impairment; MMSE = Mini-Mental State Examination; MoCA = Montreal Cognitive Assessment.
<sup>a</sup>Congestive heart failure, atrial fibrillation, ischemic heart disease, hypertension, dyslipidaemia, diabetes mellitus, cerebrovascular disease, or current tobacco use.

Based on local visual reads, 61% of scans were positive and 39% negative. Patients with visually positive versus negative scans differed slightly in demographic and clinical factors, with visually positive scans associated as expected with greater cognitive impairment (Table 1). CL were bimodally distributed across all PET scans (Fig. 1A) but unimodally distributed within visually positive and negative scans (Fig. 1B). Visually positive scans had a higher median CL (74 [interquartile range (IQR) 46, 99]) than visually negative scans (−2 [IQR −13, 12]; p<0.001, Mann-Whitney U).

**Figure 1:**
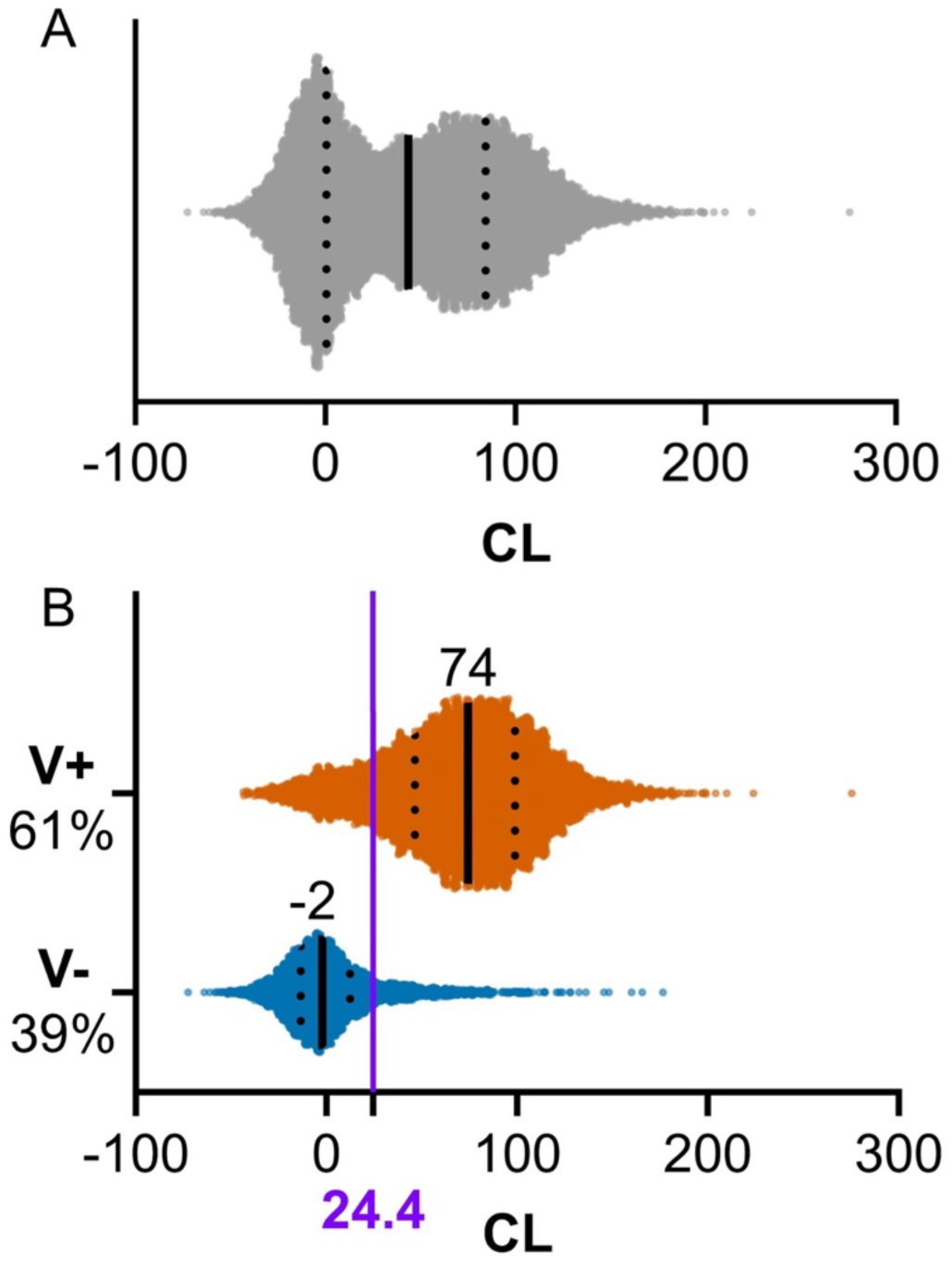
Centiloid distributions. Data points arranged into violin plots displaying the distributions of CL. **(A)** All 10,361 scans for which CL were successfully extracted. **(B)** Divided by positive vs. negative visual reads. Visually positive scans are in orange and negative in blue, with the 24.4 CL quantitative positivity threshold in purple. Violins’ vertical sizes are within-panel proportional to the number of cases. Numbers over violins and solid lines indicate medians. Dotted lines indicate 1^st^ and 3^rd^ quartiles. CL = Centiloids; MCI = mild cognitive impairment; V+ = visually positive; V-= visually negative.

### Visual read agreement with a predetermined 24.4-CL threshold

Based on our *a priori-*defined threshold of 24.4 CL, 59% of scans were quantitatively positive and 41% negative.^30^ Agreement between local visual reads and quantitative classification was 86.3% (95% CI: 85.7%, 87.0%), corresponding to a Cohen’s Kappa of 0.715 (95% CI: 0.701, 0.729; p<0.001). Overall, 5,519 [53% of total] scans were positive by both visual read and quantification (V+/Q+); 3,416 [33%] were negative by both (V-/Q-), 813 [8%] were V+/Q- and 602 [6%] were V-/Q+ (Fig. 2A). Concordantly classified scans showed a bimodal distribution of CL (median 81 CL for V+/Q+ scans and −5 CL for V-/Q- scans), while discordant scans showed a unimodal CL distribution with a median of 19 CL (Fig. 2B).

**Figure 2:**
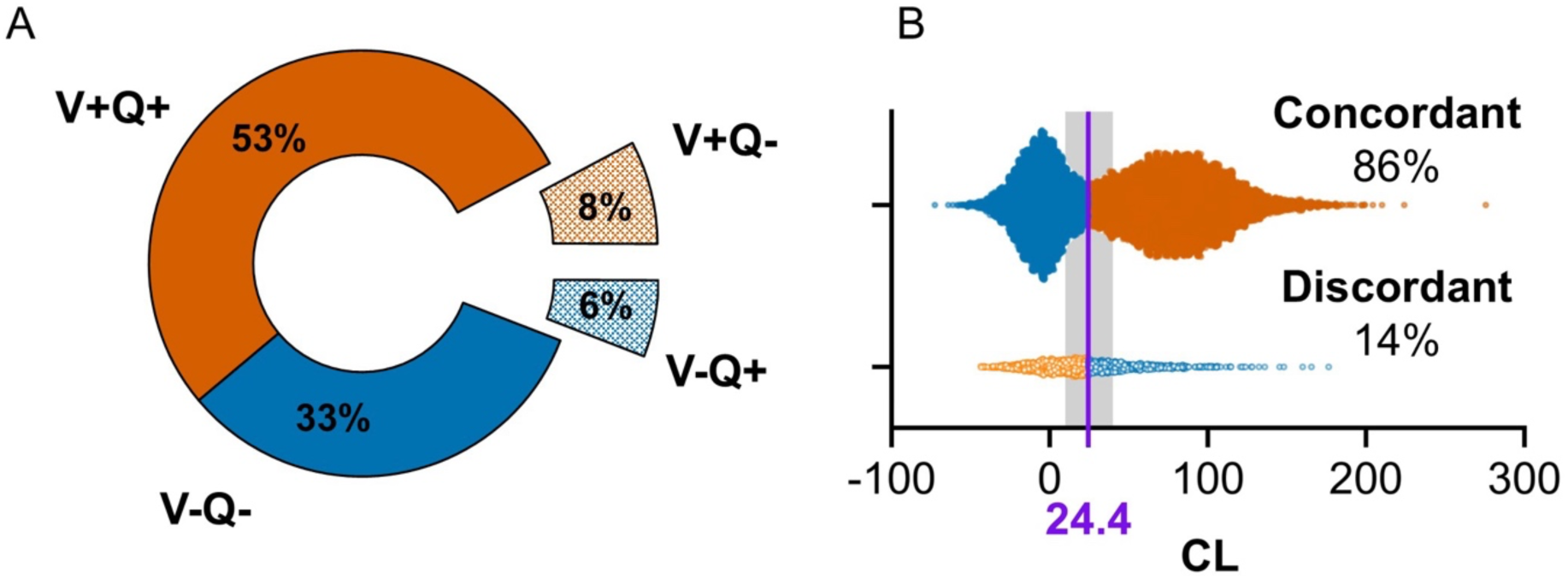
Visual-quantitative concordance. Visual-quantitative concordance based on local visual reads and the 24.4 CL quantitative positivity threshold (n=10,350). Visually positive scans are in orange and negative in blue. **(A)** Doughnut chart showing the proportions of scans by visual and quantitative classification. Concordant scans are smooth, and discordant scans are patterned and exploded. **(B)** Data points arranged in violins displaying the distribution of CL of concordant and discordant scans. Concordant scans are smooth, and discordant scans are patterned. Violins’ vertical sizes are proportional to the number of samples. The 24.4 CL threshold is marked in purple, and the 10-40 CL intermediate zone is shaded in grey. CL = Centiloids; V = visual read result; Q = quantitative result.

### Factors associated with visual-quantitative discordance

Table 2 reports associations between patient characteristics and visual-quantitative discordance. PET scans of male patients had higher rates of visual-quantitative discordance (755 [14.8%]) than scans of female patients (660 [12.6%]; p=0.001, Cramer’s V [CV]=0.032, Chi-Square). Among discordant scans, males had disproportionately more visually positive and quantitatively negative scans than females (p<0.001, CV=0.109). Self-identification as White was associated with concordance (p=0.046, CV=0.020). The radiotracers ^18^F-flutemetamol and ^18^F-florbetaben were associated with concordance (p<0.001, CV=0.038) compared to ^18^F-florbetapir, and, post-hoc, ^18^F-flutemetamol scans were more likely to be V+/Q-than the other radiotracers (p=0.035, CV=0.069). Supplementary Table 2 describes baseline characteristics and radiotracers across the four visual-quantitative combinations.

**Table 2.** Visual-quantitative concordance by patient characteristics and tracers.

|  | Concordant | Discordant | p | ES | V+Q- | V-Q+ | p | ES |
| --- | --- | --- | --- | --- | --- | --- | --- | --- |
| N (%) | 8935 (86.3) | 1415 (13.7) |  |  | 813 (57.5) | 602 (42.5) |  |  |
| Age, median (IQR) | 75 (71, 80) | 75 (70, 80) | 0.697 | 0.006 |  |  |  |  |
| Sex |  |  |  |  |  |  |  |  |
| Female (%) | 4,581 (87.4) | 660 (12.6) | 0.001 | 0.032 | 341 (51.7) | 319 (48.3) | <0.001 | 0.109 |
| Male (%) | 4,354 (85.2) | 755 (14.8) |  |  | 472 (62.5) | 283 (37.5) |  |  |
| Impairment level |  |  |  |  |  |  |  |  |
| Dementia (%) | 3,311 (85.9) | 544 (14.1) | 0.315 | 0.010 |  |  |  |  |
| MCI (%) | 5,624 (86.6) | 871 (13.4) |  |  |  |  |  |  |
| MMSE, media (IQR) | 26<br>(n=7,005) | 26<br>(n=1,101) | 0.125 | 0.029 |  |  |  |  |
| MoCA (mean ± SD) | 22 (18, 25)<br>(n=2,341) | 22 (19, 25)<br>(n=385) | 0.082 | 0.055 |  |  |  |  |
| Race |  |  |  |  |  |  |  |  |
| American Indian (%) | 21 (91.3) | 2 (8.7) | 0.487 | 0.007 |  |  |  |  |
| Alaskan Native (%) | 1 (100) | 0 (0.0) | 0.691 | 0.004 |  |  |  |  |
| Asian (%) | 154 (81.9) | 34 (18.1) | 0.075 | 0.017 |  |  |  |  |
| Black (%) | 261 (82.6) | 55 (17.4) | 0.050 | 0.019 |  |  |  |  |
| Native Hawaiian or Pacific Islander (%) | 8 (100) | 0 (0.0) | 0.260 | 0.011 |  |  |  |  |
| White (%) | 7,900 (86.6) | 1,225 (13.4) | 0.046 | 0.020 | 710 (58.0) | 515 (42.0) | 0.33 | 0.026 |
| Ethnicity |  |  |  |  |  |  |  |  |
| Hispanic or Latino (%) | 390 (86.9) | 59 (13.1) | 0.737 | 0.003 |  |  |  |  |
| Past medical history |  |  |  |  |  |  |  |  |
| Vascular Risk Factors<sup>a</sup> (%) | 6,292 (86.0) | 1,021 (14.0) | 0.183 | 0.013 |  |  |  |  |
| Tracer |  |  |  |  |  |  |  |  |
| <sup>18</sup>F-florbetapir (%) | 5,712 (85.4) | 978 (14.6) |  |  | 556 (56.9) | 422 (43.1) |  |  |
| <sup>18</sup>F-florbetaben (%) | 2,664 (87.9) | 368 (12.1) | <0.001 | 0.038 | 207 (56.3) | 161 (43.8) | 0.035 | 0.069 |
| <sup>18</sup>F-flutemetamol (%) | 559 (89.0) | 69 (11.0) |  |  | 50 (72.5) | 19 (27.5) |  |  |

For factors significantly associated with discordance, post-hoc testing of discordance type is presented.
ES denotes rank biserial correlation and Cramer's V for continuous and categorical variables, respectively. ES = effect size; IQR = Interquartile range; MCI = mild cognitive impairment; MMSE = Mini-Mental State Examination; MoCA = Montreal Cognitive Assessment.
<sup>a</sup>Congestive heart failure, atrial fibrillation, ischemic heart disease, hypertension, dyslipidaemia, diabetes, cerebrovascular disease, or current tobacco use.

We next used logistic regression to assess the associations between patient and scan factors and positive visual reads (Supplementary Table 3). In step 1 of the regression, CL alone strongly predicted positive visual reads (ROC-AUC=0.913, p<.001, n=10,349; Fig. 3A). In step 2, age, sex, impairment level (MCI/dementia), race, ethnicity, presence of at least one vascular risk factor or comorbidity, and radiotracer were added to the model, yielding a ROC-AUC of 0.915, a non-meaningful improvement compared to step 1 (ROC-AUC difference = 0.002, p for difference <0.001).

**Figure 3:**
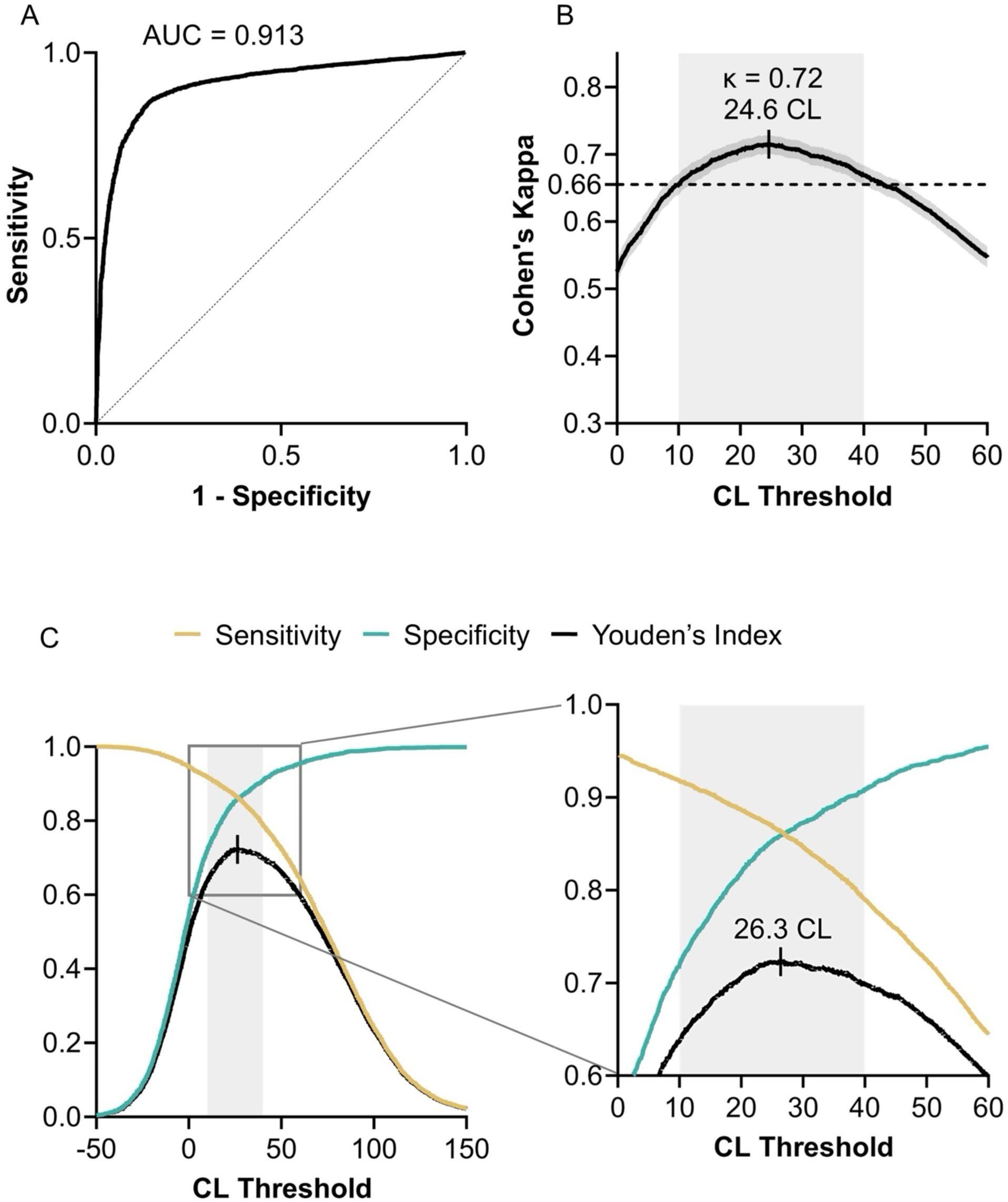
*A priori* threshold-free visual-quantitative concordance. **(A)** ROC of a logistic regression of CL as a predictor of visual read. **(B)** Cohen’s Kappa for visual-quantitative concordance across a range of CL positivity thresholds with 0.1 CL increments. Shading indicates 95% confidence interval for Cohen’s Kappa. The horizontal dotted line indicates the minimal Cohen’s Kappa within the intermediate CL range. The maximal Cohen’s Kappa is ticked. **(C)** Indices of the CL-visual read ROC and a zoom-in on the area marked with a square. The maximal Youden’s index is ticked. Across panels, the grey boxes mark the 10-40 CL intermediate range. AUC = Area under the curve; CL = Centiloids; ROC = Receiver operating characteristic curve.

### Intermediate CL zone

Quantification within the 10-40 CL intermediate zone (Supplementary Table 1) was found in 1,567 (15%) scans. Such values were more common among visually-quantitatively discordant (37%) than concordant (12%) scans, whereas CL above 40 were more common in concordant (56%) than discordant (26%) scans (p<0.001, Supplementary Fig. 1). Concordance outside the intermediate zone (89.9%, Cohen’s Kappa = 0.785) was higher than inside it (66.5%, Cohen’s Kappa = 0.331; p<0.001). Concordance above 40 CL and below 10 CL was 93.1% and 84.7%, respectively.

### Visual-quantitative agreement across CL thresholds

As studies have proposed varying CL thresholds to optimally detect Aβ positivity, we calculated Cohen’s Kappa for visual-quantitative agreement across a range of thresholds (Fig. 3B). Cohen’s Kappa exceeded 0.66 across the 10-40 CL intermediate zone, indicating strong visual-quantitative agreement across this range. The highest Cohen’s Kappa was 0.715, achieved with a threshold of 24.6 CL. This threshold and its resulting Cohen’s Kappa were nearly identical to our *a priori* 24.4-CL threshold.

Alternatively, we used receiver operating characteristic curve analysis to identify a CL threshold based on Youden’s index. This approach identified an optimal threshold of 26.3 CL, yielding 86% sensitivity and specificity (Fig. 3C).

### Quantification-supported visual reads

Local visual readers reported that quantification was used to support their visual reads in 1,605 (16%) scans. Readers in the community were not guided to use any specific threshold in interpreting the scans. Regardless of CL positivity threshold, scans that were visually read with quantification support had higher visual-quantitative agreement than scans that were visually read without it (p<0.001, permutation test; Supplementary Fig. 2). Based on the *a priori* 24.4-CL cutoff, visual-quantitative agreement in scans read with quantification (Cohen’s Kappa = 0.781, 89.5%) was significantly higher than in scans read without quantification (Cohen’s Kappa = 0.703, 85.7%; p<0.001).^33^ Peak Cohen’s Kappa values were achieved at 19.7 CL (Kappa = 0.787) for scans that used quantification support and at 24.4 CL (Kappa = 0.703) for scans that did not use quantification support. However, these optimal CL thresholds were not significantly different (p=0.069, permutation test).

### Visual read confidence

A large majority of scans were read with high confidence by the local interpreting imaging specialist (8,717, 84%), 1,497 (14%) with intermediate confidence, and 136 (1%) with low confidence. Decreasing confidence in visual reads was associated with a higher proportion of scans within the 10-40 CL zone (p<0.001, Cramer’s V [CV]=0.09, Chi-square test, Supplementary Fig. 3) and higher visual-quantitative discordance (p<0.001, CV=0.27, Fig. 4).

**Figure 4:**
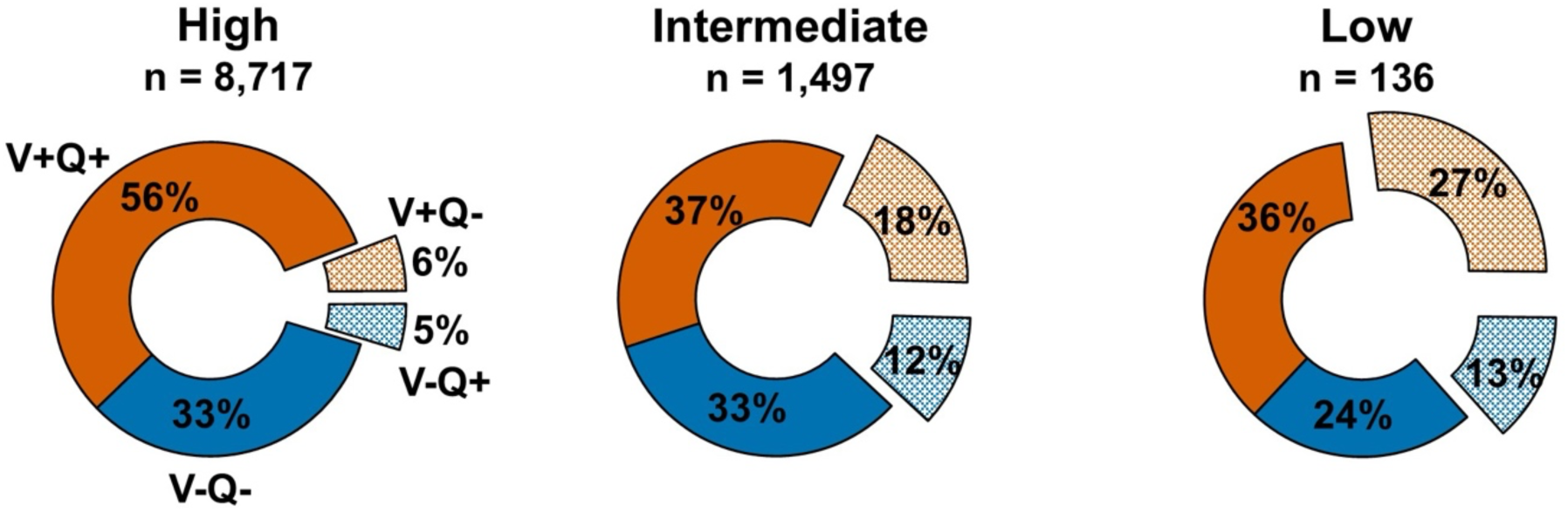
Visual read confidence. Proportions of scans by visual and quantitative classification, broken by visual read confidence. Visually positive scans are in orange and negative in blue, and concordant scans are smooth whereas discordant scans are patterned and exploded. Q = Quantification; V = Visual read.

### Association between quantification and cognitive impairment

At the whole-group level, patients with dementia had higher median CL than patients with MCI (57 [IQR 8, 91] vs. 34 [IQR −2, 79], respectively, p<0.001, rank biserial correlation [RBC]=0.15, Mann–Whitney U) as well as higher visual positivity rates (70% vs. 56%, respectively, p<0.001, Cramer’s V [CV]=0.14, Chi-square). However, when considering each visual category separately, the differences in CL levels between MCI and dementia were marginal. Specifically, visually negative scans in dementia and MCI had median CL of −3 (IQR −16, 16) and −2 (IQR −13, 12), respectively (p=0.34, RBC=0.02); visually positive scans had median CL of 76 [IQR 48, 102] and 73 [IQR 45, 97], respectively (p=0.002, RBC=0.05, Fig. 5). Within MCI, there was no significant difference between amnestic and non-amnestic MCI in CL (median=35; IQR −1, 80; n=5,262 vs. median=29; IQR −2, 78; n=1,237, respectively; p=0.18, RBC=0.03) or visual positivity rate (56% vs. 55%, respectively; p=0.48, CV=0.01).

**Figure 5:**
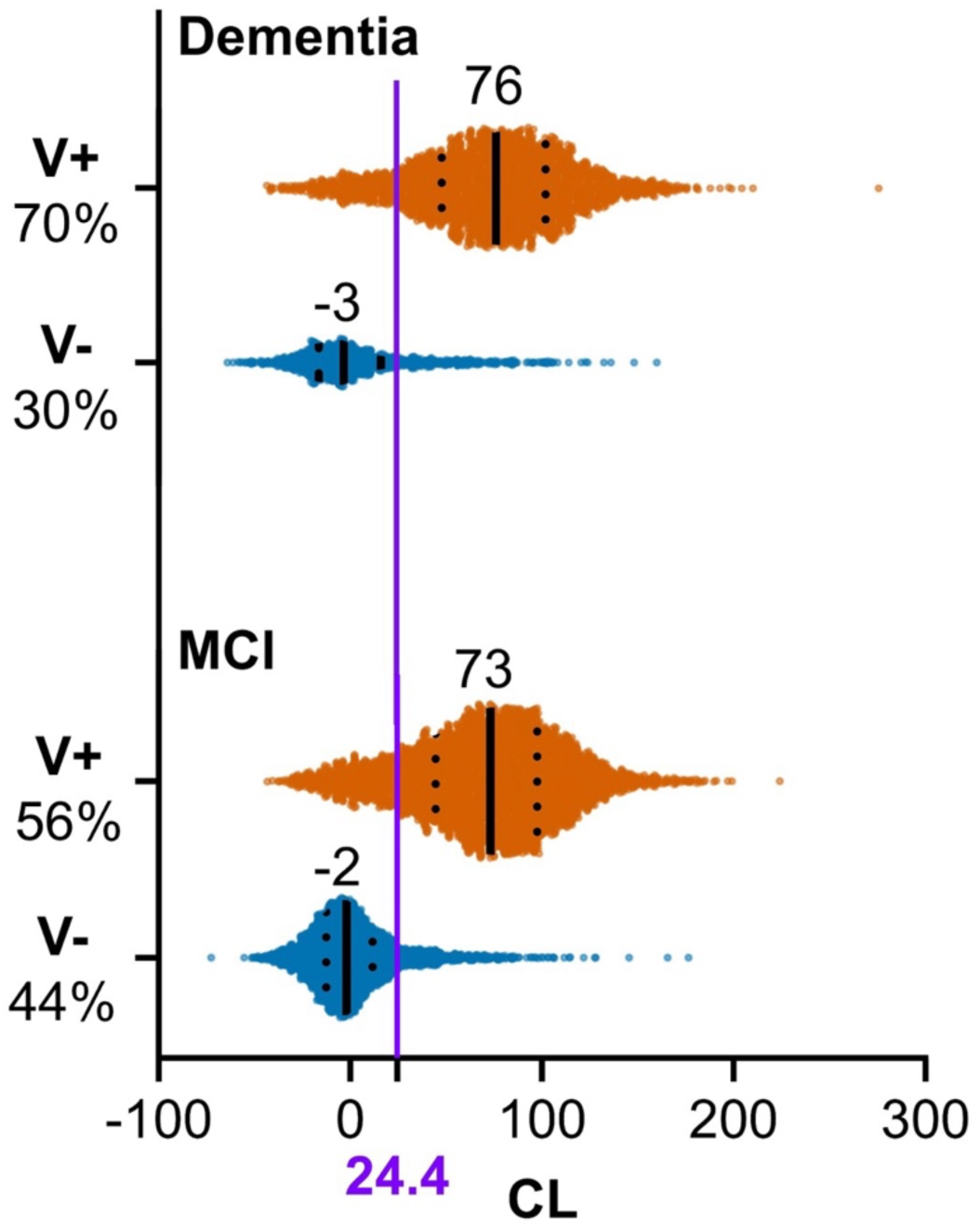
Centiloid distributions by cognitive impairment level. Data points arranged into violin plots displaying the distributions of CL divided by visual read and cognitive impairment level. Visually positive scans are in orange and negative in blue, with the 24.4 CL quantitative positivity threshold in purple. Violins’ vertical sizes are proportional to the number of samples. Numbers over violins and solid lines indicate medians. Dotted lines indicate 1^st^ and 3^rd^ quartiles. CL = Centiloids; MCI = mild cognitive impairment; V+ = visually positive; V-= visually negative.

## Discussion

Aβ-PET has an increasingly important clinical role in the diagnosis and management of patients with cognitive decline, especially with the recent approval of Aβ-targeting monoclonal antibodies, which require confirmation of brain amyloidosis via PET or CSF studies for treatment eligibility.^36,37^ Aβ-PET interpretation in the clinic is based on a dichotomous visual read performed by trained imaging specialists. Previous studies validating the reliability of visual reads used standardized PET acquisition and processing and experienced imaging specialists, and thus require replication under the much less controlled environment of real-world clinical practice. In this study, we leveraged the large and heterogeneous Aβ-PET dataset collected in IDEAS to evaluate the concordance between community visual reads and CL quantification performed centrally. Overall, we found 86% agreement between quantitative classification of scans and local visual reads, corresponding with an excellent Cohen’s Kappa of 0.715. Importantly, high rates of agreement between quantification and visual reads were found across a range of possible quantitative thresholds, supporting the robustness of our findings. Our study also demonstrates the feasibility of CL quantification in community-acquired Aβ-PET scans, with derived CL values and distributions closely matching those described in the research setting and clinical trials.^38–42^ Overall, our results strongly support the validity of Aβ-PET as used in real-world clinical settings.

The visual-quantitative agreement we found for Aβ-PET is well within the range of other neuroimaging modalities used in the workup of neurodegenerative conditions. In ^18^F-fludeoxyglucose PET, the agreement between visual reads and automated quantitative analysis for determining AD pattern hypometabolism is 62.3%-84.4%, depending on reader expertise and clinical severity.^43^ For dopamine transporter single-photon emission computed tomography, one study found 91% agreement,^44^ and another found Cohen’s Kappa of 0.66 to 0.72,^45^ for concordance between expert reads and quantification of striatal radiotracer binding. For MRI-measured brain atrophy, correlation coefficients between automated quantification and visual scales range from 0.20 to 0.72.^46–48^ These agreement rates are largely derived from research-level data, which are expected to yield higher rates than the real-world cohort we analyzed. Studies evaluating agreement in Aβ-PET in research settings, employing panels of expert visual readers and quantification extracted using various methods and cutoffs, reported Cohen’s Kappa values of 0.85-0.92 and percent agreement of 88.9-100%.^24,25,49^ Thus, whereas agreement may approach perfection under certain optimal conditions, Aβ-PET real-world concordance lagged only slightly behind. Real-world results similar to ours were reported by the AMYPAD-DPMS study, which found 92.3% agreement (Cohen’s Kappa of 0.85) between local readers and CL status.^50^ The slightly lower real-world concordance is likely attributable to a more heterogeneous cohort, less stringent PET acquisition, less experienced visual readers, and less robust CL extraction. In our study, discordance between visual reads and quantification was driven to a large extent by intermediate scans, characterized by quantification within the 10-40 CL range and associated with lower confidence in the visual read. It is increasingly recognized that AD biomarker results around the positivity cutoff should be classified as intermediate, and our findings can help clinicians identify patients more likely to have such Aβ-PET results.^31^

Quantitative Aβ-PET analysis can potentially be used to guide AD disease-modifying treatments beyond visual reads. In clinical trials, Aβ-PET quantification is used for patient selection, ^41,51–53^ as an outcome measure to evaluate target engagement,^51,53–56^ and even to discontinue or adjust Aβ-lowering therapy after a predefined effect has been reached.^41,53^ In addition, the continuous nature of quantitative analysis allows the identification of intermediate scans with borderline signal intensity. The optimal implementation of all these potential clinical uses of Aβ-PET would be greatly enhanced by quantification of Aβ-PET in real-world settings. The current study’s MRI-free CL extraction from clinical Aβ-PET scans and their correlation with visual reads support the feasibility of large-scale quantification in clinical practice, which could improve access to advanced Aβ-PET applications. In IDEAS, image quantification was used as an optional adjunct to visual reads in interpreting 16% of scans in the imaging dataset. Visual reads that incorporated local quantification showed higher agreement with our *a priori*-defined CL threshold of 24.4 CL (a threshold that was not known to the imaging specialists in the study). Optimal agreement between visual reads and quantification occurred at a slightly lower threshold of 19.7 CL, suggesting that quantification may enhance the sensitivity of visual reads. Future studies are needed to specifically evaluate the impact of quantification on clinical decision-making.

We found a CL distribution that was largely consistent with previous reports from highly controlled research settings and clinical trials, further supporting the validity of quantification of real-world clinical scans. CL displayed a bimodal distribution consisting of a mix of two unimodal distributions from visually positive and visually negative scans, as previously described in research settings that included mixed MCI and dementia cohorts.^38,39^ In our sample, the median CL of visually negative scans was −2, very close to the young controls 0 anchor of the CL scale and similar to previous cohort.^20,39^ Median CL in visually positive scans was 74, lower than the 100 CL anchor defined for patients with mild-to-moderate dementia,^20^ and slightly lower than in clinical trials recruiting Aβ-positive MCI and mild dementia patients, where mean baseline CL ranged from 75 to 103.^39–42^ We found a higher median CL in dementia than MCI, but this was largely driven by a higher rate of visually positive scans in dementia. Within visually positive scans, the median CL of MCI (73) was similar to dementia (76). Previous studies found higher mean CL in MCI (75 and 81) and in dementia (94 and 90) among Aβ-PET positive participants.^39,57^ It is possible that the slightly lower CL in our cohort is driven by the more heterogeneous population, with non-AD pathology contributing to cognitive decline even in visually positive patients with dementia. Our cohort included a high rate of patients with vascular risk factors and comorbidities, who were excluded from previous studies.

PET-to-autopsy studies suggest that a moderate degree of Aβ accumulation is required for reliable detection by Aβ-PET.^30,58^ Our literature search found a range of proposed CL positivity cutoffs, which generally fell within a 10-40 CL range. Higher cutoffs are associated with greater Aβ burden at autopsy and with higher specificity but lower sensitivity for detecting Aβ aggregation.^30,59^ Identification of such intermediate zones of AD biomarker results was recently suggested in the updated Alzheimer’s Association AD diagnostic criteria.^31^ The lower visual read confidence in scans within the intermediate zone implies another characteristic common for both methods, as readers more often identified scans in the intermediate CL range as challenging to interpret visually. Overall, it seems likely that a proportion of the scans that are intermediate visually and quantitatively reflect the minimum burden of Aβ accumulation that can be detected by PET.

Limitations of this study should be considered. The study did not include any neuropathological data and thus cannot directly compare Aβ-PET with autopsy as the standard of truth. Thus, we must rely on the internal and external consistency of the results to assess the performance of the test. CL were extracted using an MRI-free pipeline (rPOP), which might differ from the standard MRI-based method. However, rPOP-derived CL showed very high correlation (R^2^=0.95) with CL derived from an MRI-based pipeline.^29^ Thus, it is unlikely that our results have been significantly affected. Some centers opted not to submit images and were thus not included in the analysis. It is possible that those were centers with less emphasis on research or PET imaging, or have more heterogenous patients, resulting in lower quality of scan acquisition and interpretation. The lack of meaningful difference in patient characteristics between scans for which CL were extracted and the entire IDEAS dataset indirectly suggests otherwise but does not rule out that option (Supplementary Table 7). On the other hand, our uniquely large database, derived from many clinical centers from across the USA, greatly strengthens the generalizability of our results.

In conclusion, in a large heterogeneous dataset of real-world Aβ-PET scans, we found high concordance between local visual reads and image quantification. Quantification of real-world PET scans shows comparable CL values and distributions to those reported in observational research studies and clinical trials. Our findings support the validity of Aβ-PET as acquired and interpreted in real-world clinical settings.

## Supplementary material

Supplementary material is available at *Brain* online.

## Data Availability

All data presented in this manuscript (clinical, demographic, and imaging variables, including CL values) are available through the Global Alzheimer’s Association Interactive Network (https://www.gaaindata.org/partner/IDEAS). Raw and processed PET images are also available through the IDA at Laboratory of Neuro Imaging (https://ida.loni.usc.edu/login).

