## Supplemental Data for "Concordance between amyloid-PET quantification and real-world visual reads: results from IDEAS"

**Supplementary Table 1 CL positivity thresholds suggested in the literature.**

| CL Threshold | Tracer | Standard of truth | Reference |
|---|---|---|---|
| 10 | FBB, PiB | Pathology | 1 |
| 12 | FBP, FTM | CSF AD biomarkers | 2 |
| 12.2 | PiB | Pathology | 3 |
| 13.5 | FBB | Cognitively normal young controls | 4 |
| 16 | FTM | Visual reads | 5 |
| 17 | FTM | Visual reads | 6 |
| 19 | PiB | Longitudinal progression | 7 |
| 19 | FBB | Pathology | 8 |
| 21 | FBB | Visual reads | 9 |
| 24.4 | PiB | Pathology | 3 |
| 26 | FTM | Longitudinal progression | 10 |
| 26 | FBB, PiB | Visual reads | 1 |
| 30 | FBP, FTM | CSF AD biomarkers | 2 |
| 35.7 | FBB | Clinical diagnosis and visual reads | 4 |
| 49.4 | FBB, PiB | Pathology | 1 |

AD = Alzheimer's disease; CL = Centiloids; CSF = Cerebrospinal fluid; FBB = $^{18}$F-florbetaben; FBP = $^{18}$F-florbetapir; FTM = $^{18}$F-flutemetamol; PiB = Pittsburgh compound B.

**Supplementary Table 2 Patient characteristics and tracers across the four visual-quantitative combinations.**

| | V+Q+ | V-Q- | V+Q- | V-Q+ |
|---|---|---|---|---|
| N (%) | 5519 (53.3) | 3416 (33.0) | 813 (7.9) | 602 (5.8) |
| Age, median (IQR) | 76 (72, 81) | 74 (70, 79) | 75 (70, 80) | 75 (71, 81) |
| **Sex** | | | | |
| Female (%) | 2976 (56.8) | 1605 (30.6) | 341 (6.5) | 319 (6.1) |
| Male (%) | 2543 (49.8) | 1811 (35.4) | 472 (9.2) | 283 (5.5) |
| **Impairment level** | | | | |
| Dementia (%) | 2385 (61.9) | 926 (24.0) | 316 (8.2) | 228 (5.9) |
| MCI (%) | 3134 (48.3) | 2490 (38.3) | 497 (7.7) | 374 (5.8) |
| MMSE, median (IQR) | 25 (21, 27) (n=4272) | 27 (24, 29) (n=2733) | 26 (23, 28) (n=611) | 26 (22, 28) (n=490) |
| MoCA (mean ± SD) | 21 (17, 24) (n=1507) | 23 (20, 26) (n=834) | 22 (19, 25) (n=246) | 22 (19, 25) (n=139) |
| **Race and ethnicity** | | | | |
| American Indian (%) | 10 (43.5) | 11 (47.8) | 1 (4.3) | 1 (4.3) |
| Alaskan Native (%) | 1 (100) | 0 (0.0) | 0 (0.0) | 0 (0.0) |
| Asian (%) | 67 (35.6) | 87 (46.3) | 21 (11.2) | 13 (6.9) |
| Black (%) | 148 (46.8) | 113 (35.8) | 26 (8.2) | 29 (9.2) |
| Native Hawaiian or Pacific Islander (%) | 3 (37.5) | 5 (62.5) | 0 (0) | 0 (0) |
| White (%) | 4,980 (54.6) | 2,920 (32.0) | 710 (7.8) | 515 (5.6) |
| **Ethnicity** | | | | |
| Hispanic or Latino (%) | 214 (47.7) | 176 (39.2) | 37 (8.2) | 22 (4.9) |
| **Past medical history** | | | | |
| Vascular Risk Factors[a] (%) | 3858 (52.8) | 2434 (33.3) | 595 (8.1) | 426 (5.8) |
| **Tracer** | | | | |
| $^{18}$F-florbetapir (%) | 3522 (52.6) | 2190 (32.7) | 556 (8.3) | 422 (6.3) |
| $^{18}$F-florbetaben (%) | 1636 (54.0) | 1028 (33.9) | 207 (6.8) | 161 (5.3) |
| $^{18}$F-flutemetamol (%) | 361 (57.5) | 198 (31.5) | 50 (8.0) | 19 (3.0) |

IQR = Interquartile range; MCI = mild cognitive impairment; MMSE = Mini-Mental State Examination; MoCA = Montreal Cognitive Assessment.
[a]Congestive heart failure, atrial fibrillation, ischemic heart disease, hypertension, dyslipidaemia, diabetes, cerebrovascular disease, or current tobacco use.

**Supplementary Table 3 Logistic regression predicting visual positivity.**

| Predictor | Estimate | OR | OR 95%CI | | p |
|---|---|---|---|---|---|
| **Step 1** | | | | | |
| Intercept | -1.240 | 0.289 | 0.269 | 0.311 | <0.001 |
| CL | 0.052 | 1.054 | 1.052 | 1.056 | <0.001 |
| AUC-ROC = 0.9133 | | | | | |
| **Step 2** | | | | | |
| Intercept | -3.082 | 0.046 | 0.022 | 0.096 | <0.001 |
| CL | 0.052 | 1.053 | 1.051 | 1.055 | <0.001 |
| Age (y) | 0.020 | 1.020 | 1.011 | 1.030 | <0.001 |
| Female – Male | -0.039 | 0.962 | 0.856 | 1.081 | 0.515 |
| Dementia – MCI | 0.428 | 1.534 | 1.353 | 1.738 | <0.001 |
| American Indian | -0.765 | 0.465 | 0.125 | 1.731 | 0.254 |
| Alaskan Native | 10.173 | 26,173 | 0.000 | >999 | 0.959 |
| Asian | 0.154 | 1.167 | 0.736 | 1.850 | 0.511 |
| Black | 0.009 | 1.009 | 0.681 | 1.494 | 0.965 |
| Native Hawaiian or Pacific Islander | -0.256 | 0.774 | 0.112 | 5.333 | 0.795 |
| White | 0.333 | 1.395 | 1.104 | 1.762 | 0.005 |
| Hispanic or Latino | 0.073 | 1.076 | 0.807 | 1.435 | 0.618 |
| Vascular Risk Factor/Comorbidity | -0.017 | 0.983 | 0.864 | 1.118 | 0.797 |
| FBB – FBP | -0.214 | 0.807 | 0.709 | 0.919 | 0.001 |
| FTM – FBP | 0.184 | 1.202 | 0.946 | 1.527 | 0.131 |
| AUC-ROC = 0.9154 | | | | | |

A logistic regression model predicting visual positivity over negativity. AUC-ROC = area under the receiver operating characteristic curve; CI = confidence interval; CL = Centiloids; FBB = $^{18}$F-florbetaben; FBP = $^{18}$F-florbetapir; FTM = $^{18}$F-flutemetamol; OR = odds ratio.

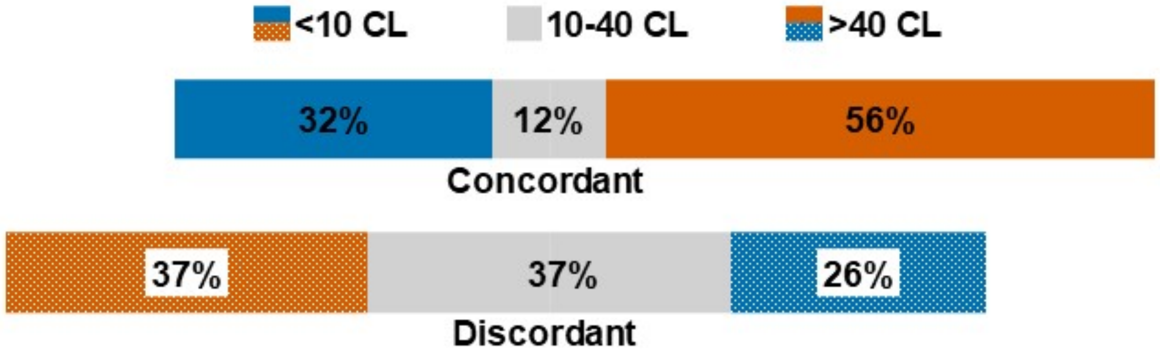


**Supplementary Figure 1: Proportions of scans by CL group and visual-quantitative concordance.** CL = Centiloids.

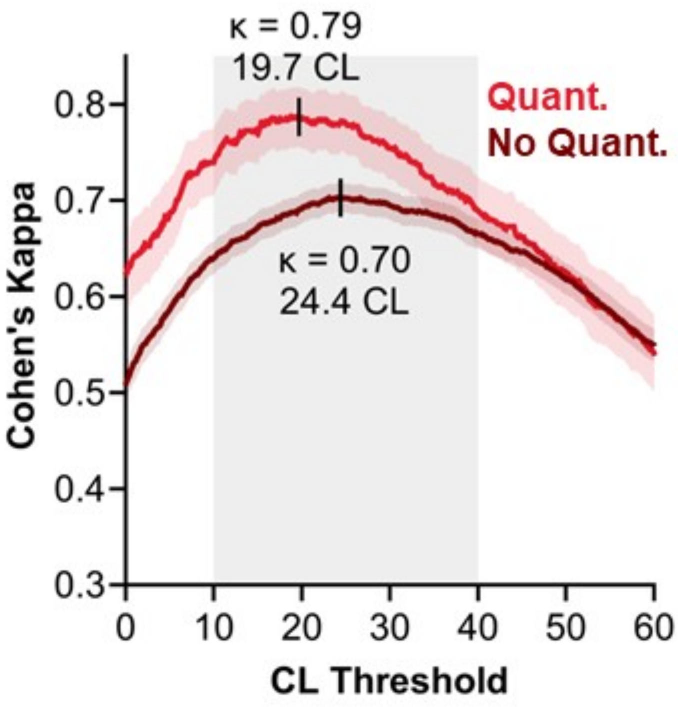


**Supplementary Figure 2: Visual-quantitative concordance across thresholds by use of quantification to support visual reads.** Cohen's Kappa for visual-quantitative concordance across a range of quantitative positivity thresholds with 0.1 CL increments, divided by the use of quantification to support visual reads. Shadings indicate 95% confidence interval for Cohen's Kappa. Maximal Cohen's kappa and corresponding threshold for each category are ticked in black. The grey box marks the 10-40 CL borderline range. CL = Centiloids. Quant. = Use of quantification to support visual reads.

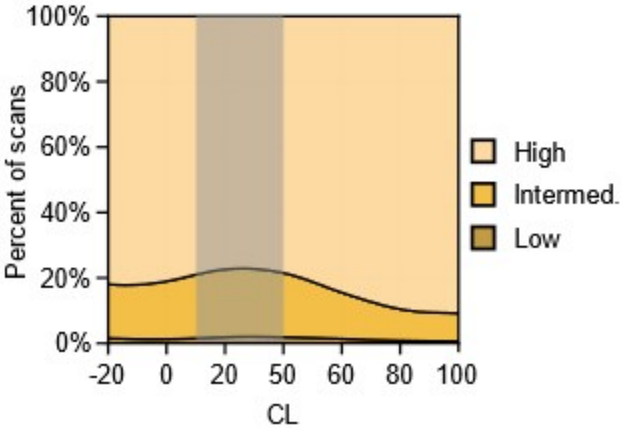


**Supplementary Figure 3:** The proportion of scans with low, intermediate, and high visual read confidence ratings as a function of CL, with a Gaussian kernel smoothing. CL = Centiloids; Interm. = Intermediate; Prop. = Proportion.

### Supplementary statistical methods

A permutation testing procedure was used to assess differences in (1) the strength of visual-quantitative agreement, and (2) optimal CL thresholds between quantification- and non-quantification-supported visual reads. First, we identified the CL thresholds that maximized Cohen's kappa across scans in each of these categories, testing CL from 0 to 60, as described above. We then shuffled the labels of which scans had been read with, versus without, quantitative support and recalculated maximum kappas and corresponding CL thresholds in the shuffled dataset, repeating this process 5,000 times independently to obtain a null distribution. Finally, we calculated empirical p-values to determine whether quantification- and non-quantification-supported visual reads differed significantly in the level of visual-quantitative agreement (maximum kappa across the CL range examined) or optimal CL threshold. Specifically, we applied the formula $p = \frac{r+1}{n+1}$, where r is the number of shuffled iterations for which the absolute value of the difference between maximum kappas or optimal CL thresholds, respectively, in the two groups equaled or exceeded the absolute value of their difference in the observed (unshuffled) data, and n is the total number of shuffled permutations.

### Supplementary PET processing methods

PET scans were uploaded by the acquiring facilities to the American College of Radiology, then transferred to the Laboratory of Neuroimaging (LONI), and downloaded by the UCSF team to be centrally processed and quantified.

*Processing*

The raw PET data, initially available in DICOM format, underwent a series of processing steps as outlined in Table 1. DICOM-to-NIFTI conversion was performed for each session, resulting in a single-frame file for most sessions. Some sessions yielded multiple frames, in which case the frames were realigned using rigid-body transformation and averaged. Then, the PET scans were processed using an approach specifically designed for this dataset: *robust PET-Only Processing* (rPOP (*1*)). In short, this involved warping the scans to the Montreal Neurological Institute (MNI) template space through a mix of affine and non-linear transformations, utilizing SPM12's Old Normalization tool (*2*) and a collection of nine PET templates in MNI space representing negative, positive, and intermediate scans for each radiotracer.

To ensure data harmonization, the image resolution was estimated directly from the PET images themselves, utilizing AFNI's 3dFWHMx function. For images with an estimated resolution below 10mm full width at half maximum (FWHM), a Gaussian filter was applied to achieve an effective resolution of 10mm FWHM. Finally, standardized uptake value ratios (SUVr) maps were generated, employing whole cerebellar masks from the Global Alzheimer's Association Interactive Network (GAAIN) Centiloid project as reference (*3*).

| **Process name** | **Title Link** | **Program/Package** |
|---|---|---|
| Convert from DICOM series to NIFTI file | Conv | dicm2nii |
| *In case of multi-frame DICOM series*: realign frames using rigid-body transformation and average | Realign, Avg | SPM12, spm_realign |

| | | |
|---|---|---|
| Non-rigid transform (warp) to standard image space and voxel size | Non-Rigid Reg to Std Img Vox Size | rPOP |
| Estimate image resolution | - | AFNI, 3dFWHMx |
| Smooth to effective resolution of 10 mm FWHM | Uniform Res | SPM, spm_smooth |
| Compute SUVr images (reference: whole cerebellum) | Inten Norm | NumPy |

**Supplementary Table 4. List of steps in chronological order applied to process PET data with the corresponding program and/or package name used for each process.**

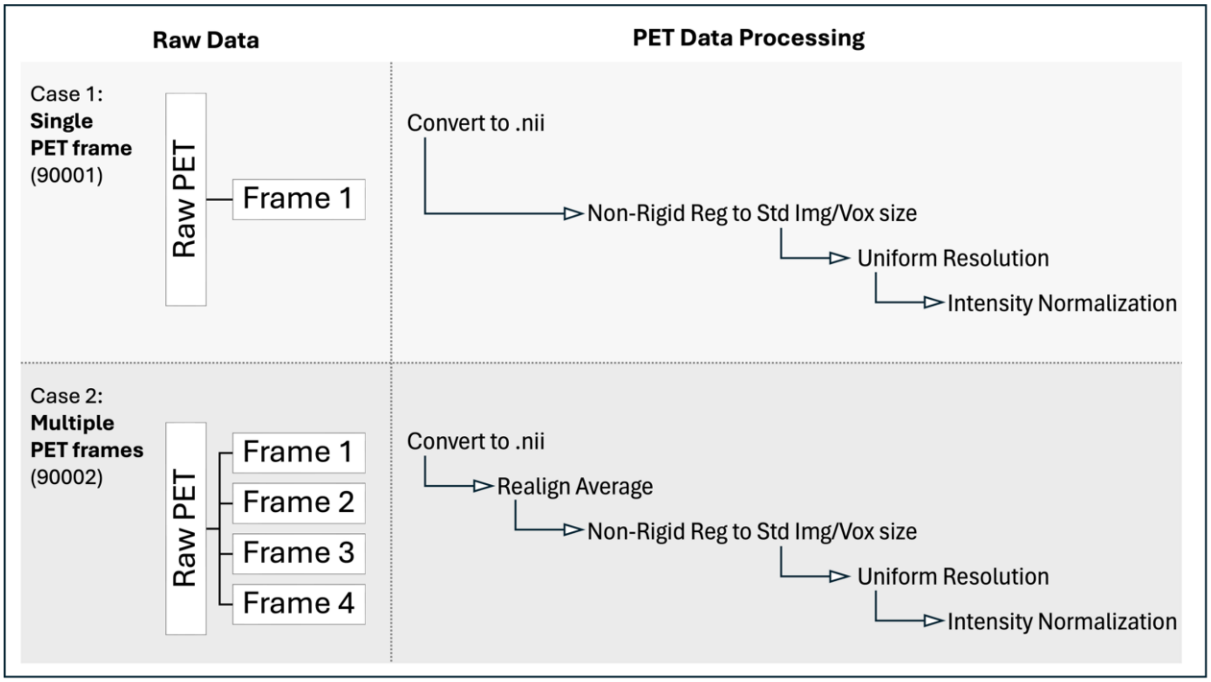


**Supplementary Figure 4. Example of the processing applied to the two cases with different number of PET frames available.**

*LONI image terms*

In addition to the raw DICOM files, three types of processed images are available in the dataset, each of which is detailed in Table 2. These files can be located by utilizing the 'Advanced Search' function and selecting the 'Processed' option under the 'Image Types' section of the search page. Type 1 images maintain the same image size, voxel dimensions, and spatial orientation as the original PET image data. Type 2 images undergo transformation to MNI space, resulting in dimensions of 101 x 116 x 96 with 2 mm isotropic voxels. Type 3 images retain the spatial orientation of Type 2 images.

The term "<Tracer>" in the label signifies the final tracer ([$^{18}$F]Florbetapir, [$^{18}$F]Florbetaben, or [$^{18}$F]Flutemetamol) identified as the true tracer injected, which serves as crucial information for selecting the appropriate CL conversion equation. The tracer designation is determined through an examination of available information provided by sites on two different forms.

**Supplementary Table 5. LONI image description terms for each available processed image type.**

| **DICOM Frames** | **LONI Image Description** | **Image Type** |
|---|---|---|
| Single | PET <Tracer> Conv | 1 |
| | PET <Tracer> Conv, Non-Rigid Reg to Std Img Vox Size | 2 |
| | PET <Tracer> Conv, Non-Rigid Reg to Std Img Vox Size, Uniform Res, Inten Norm | 3 |
| Multiple | PET <Tracer> Conv, Realign, Avg | 1 |
| | PET <Tracer> Conv, Realign, Avg, Non-Rigid Reg to Std Img Vox Size | 2 |
| | PET <Tracer> Conv, Realign, Avg, Non-Rigid Reg to Std Img Vox Size, Uniform Res, Inten Norm | 3 |

*Quality Control (QC)*

All scans underwent standardized visual QC, which was performed on Type 2 images (i.e., images in standard space and voxel size, prior to smoothing and intensity normalization, see above). Each scan was reviewed by two independent raters using a standardized process displayed in Figure 2.

This visual QC aimed to identify 3 types of issues:

i) Non-fixable issue with the PET scan itself (e.g. reconstruction or attenuation correction issue), resulting in the scan being excluded from analyses.

ii) Issue with the quality of the warping (non-rigid registration) to template, resulting in poor placement of the Centiloid/GAAIN Volume of Interest (VOI) masks. For these scans, Step 1 images were manually realigned before re-running the warping procedure. Scans that were not fixed by this procedure were excluded from analyses.

iii) Issue related to major anatomical lesion that would impact PET quantification (e.g., infarct, major ventriculomegaly). After visual inspection, some scans were excluded, while others, mainly with unilateral lesions, were included in the final dataset, but flagged for specific Centiloid calculation (see below).

Lastly, we used a quantitative approach to quantify the potential impact of field-of-view cuts which impact the ability to extract robust signal from either the reference or the cortical Volume of Interest (VOI). Scans with missing voxels in more than 10% voxels in either VOI were excluded from the dataset.

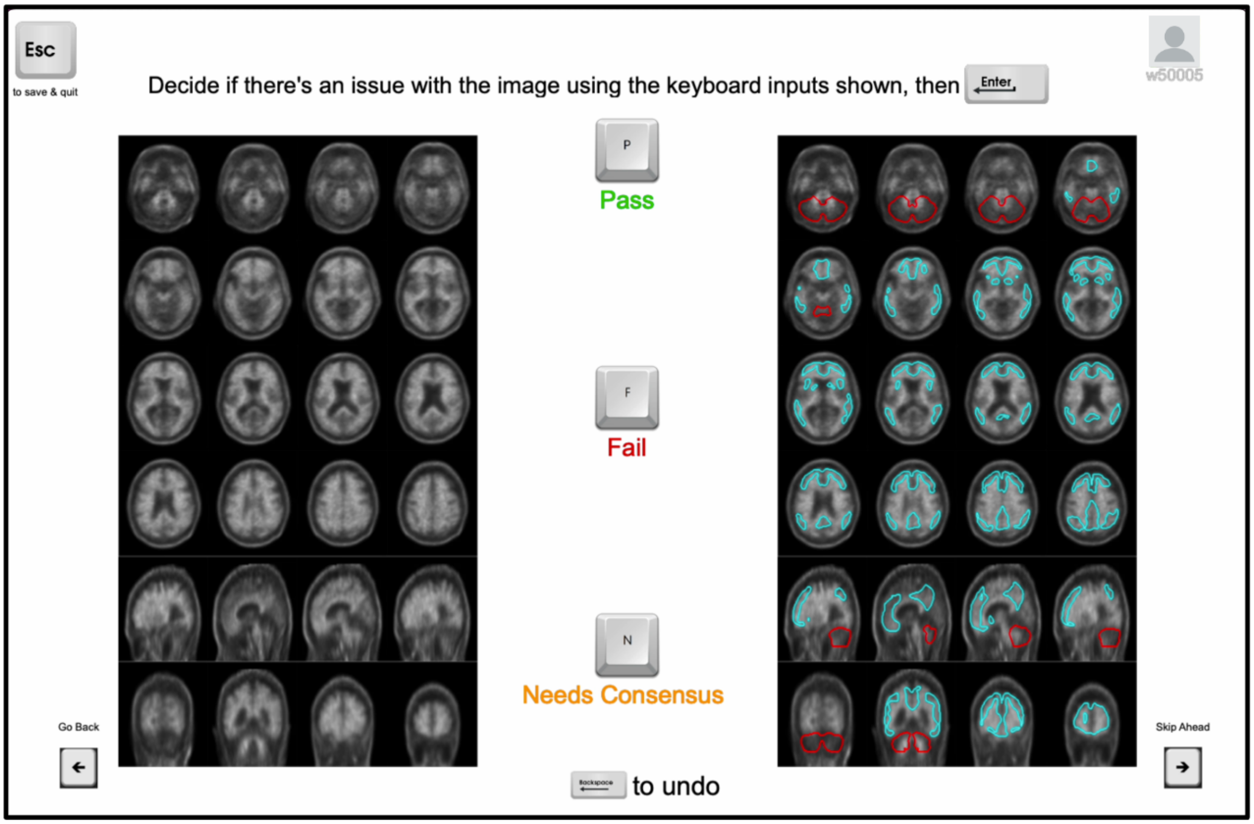


**Supplementary Figure 5. Screenshot of the visual QC program used to review each scan. Code used to create these multi-slice images is available on github: https://github.com/UCSFMemoryAndAging/MAC-PET/tree/main/IDEAS_QC**

Scans available for download passed either the initial (untouched) or second (re-processed or adapted) round of QC, and did not have major field of view cuts (final n=10,361 patients). This includes scans of participants with unilateral anatomical lesions in the reference or target regions (n=225). For these scans, SUVr values used to compute CL units were adapted, meaning that one or two VOIs included only the non-affected hemisphere.

*Centiloid calculation*

Centiloid (CL) is the standardized quantitative amyloid measure intended to facilitate comparison across different amyloid PET tracers. See Klunk et al. (*4*) for more information on the CL scale.

Proposed MRI-free pipeline uses two Volumes of Interest (VOI) to extract mean PET counts to calculate CL: the cortical and whole cerebellar masks from the GAAIN Centiloid project (see Figure 3). Corresponding masks can be found on http://www.gaain.org/centiloid-project, under "Calibration of the standard PIB method", in the Centiloid_Std_VOI.zip file.

These two values served as the target and reference regions for computing SUVr from the warped smoothed images. Subsequently, these SUVr values were converted to CL units utilizing tracer-specific equations (*1*) that had been previously validated, as detailed in Table 3.

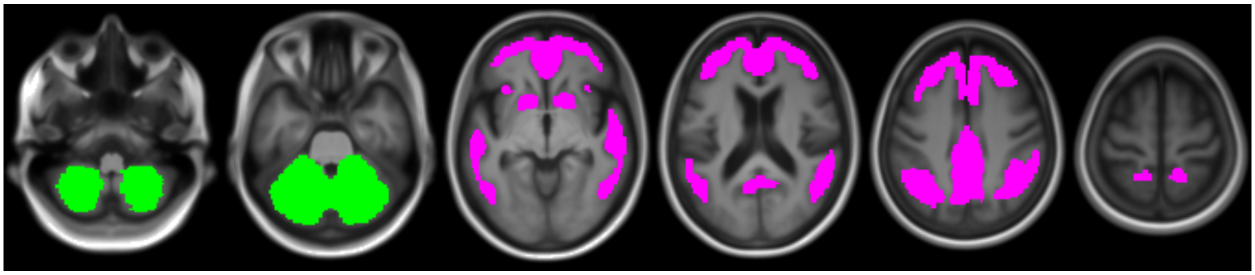

**Supplementary Figure 6. Reference (whole cerebellum, green) and target (cortical region, pink) VOIs overlaid on MRI template.**

**Supplementary Table 6. Centiloid conversion equations for cortical SUVr (ref: whole cerebellum).**

| **Tracer** | **Equation** |
|---|---|
| [$^{18}$F]Florbetapir | CL = (189.9 x Cortical_SUVr) – 211.1 |
| [$^{18}$F]Florbetaben | CL = (160.7 x Cortical_SUVr) – 169.2 |
| [$^{18}$F]Flutemetamol | CL = (127.6 x Cortical_SUVr) – 136.2 |

*Final sample*

**Supplementary Table 7.** The main demographic and clinical characteristics of the 10,361 patients with available Centiloid quantification, compared to the full IDEAS sample.

| | *Patients with CL quantification* N=10,361 | *Full IDEAS sample* N=18,293 |
|---|---|---|
| Age: median [Q1, Q3] | 75 [71,80] | 75 [71,80] |
| Gender | | |
| *Men* | 49.3% | 48.7% |
| *Women* | 50.6% | 51.3% |
| Race* | | |
| *Asian* | 1.8% | 1.8% |
| *Black* | 3.0% | 3.5% |
| *White* | 88.1% | 86.7% |
| Ethnicity: %Hispanic or Latino | 4.3% | 4.5% |
| Education | | |
| *No formal education or Grade School* | 2.8% | 3.0% |
| *Some high school* | 4.0% | 4.1% |
| *High school (including equivalency)* | 25.3% | 26.3% |
| *Some college or associate degree* | 24.1% | 23.3% |

| | | |
|---|---|---|
| *Bachelor's degree* | 23.3% | 23.3% |
| *Master's degree* | 12.3% | 11.9% |
| *Doctorate* | 8.3% | 8.2% |
| Family History of Dementia | | |
| *Yes* | 23.7% | 24.2% |
| *No* | 76.3% | 75.8% |
| Level of Clinical Impairment | | |
| *MCI* | 62.7% | 60.5% |
| *Dementia* | 37.3% | 39.5% |
| MMSE: median [Q1, Q3] | 26 [22, 28] | 26 [22, 28] |
| % Taking Cholinesterase Inhibitors at Baseline | 36.4% | 36.5% |
| Amyloid-PET result (local read) | | |
| *Positive* | 60.7% | 60.9% |
| *Negative* | 39.2% | 39.0% |
| *Uninterpretable/Technically inadequate study* | 0.1% | 0.1% |

* only indicating groups with >1% representation in the whole sample

2. SPM12; Wellcome Department of Imaging Neuroscience, Institute of Neurology, London, UK.

3. GAAIN, http://www.gaain.org/centiloid-project.

4. Klunk WE, Koeppe RA, Price JC, et al. *The Centiloid Project: standardizing quantitative amyloid plaque estimation by PET*. Alzheimers Dement. 2015;11:1-15.e1-4.
